# Prevalence of Common Respiratory Viruses in Children: Insights from Post-Pandemic Surveillance

**DOI:** 10.1101/2024.09.12.24313530

**Authors:** Constance Adu-Gyamfi, Jesse Addo Asamoah, James Opoku Frimpong, Richard Larbi, Richard Owusu Ansah, Sherihane Naa Ayeley Aryeetey, Richmond Gorman, Henry Kyeremateng Acheampong, Emmanuella Nyarko-Afriyie, Manuella Hayford, Henrietta Dede Tetteh, Kwadwo Boampong, Veronica Barnor, Peter K. Brenya, Frederick Ayensu, NK. Ayisi-Boateng, Philip El-Duah, Christian Drosten, Richard Odame Phillips, Augustina Angelina Sylverken, Michael Owusu

## Abstract

**Introduction:** The COVID-19 pandemic has significantly affected healthcare systems worldwide, impacting the occurrence and management of respiratory illnesses. This has also influenced respiratory infections’ role in childhood mortality. Surveillance of common respiratory viruses in Ghana is limited, making it crucial to assess the prevalence of respiratory viral infections, particularly in children, in the post-pandemic era. This study provides data on the prevalence of respiratory viruses and the associated risk factors in children aged 5 or younger in an urban paediatric hospital setting.

**Methods:** The study was a cross-sectional study with a convenience sampling method, conducted in four health facilities: Asokwa Children’s Hospital, HopeXchange Medical Centre, University Health Services-KNUST, and Kumasi South Hospital in Kumasi, Ghana, between August 2022 and June 2023. Recruitment was not done in parallel in each hospital. Oropharyngeal swabs were collected from children ≤ 5 years old and screened by RT-qPCR for common respiratory viruses.

**Results:** Out of the 303 patients enrolled in the study, 165 (54.4%) were male, and 122 (40.3%) were aged from 13 to 36 months. The median age of the patients was 19 months. The most common symptoms reported were cough (87.0%), runny nose (87.0%), and fever (72.0%). Respiratory viruses were detected in 100 (33.0%) of the samples, with 36 (12.0%) testing positive for Human metapneumovirus (HMPV), 27 (8.9%) for RSV, and 20 (6.6%) for Human Adenovirus (HAdV). In 8.0% of the cases, multiple viruses were detected, with HAdV being the most common (75.0%). Children under 6 months (AOR: 4.81, 95% CI: 1.20-24.60) had a higher risk of RSV detection compared to children aged 37 to 60 months. Furthermore, it was found that caregivers with tertiary education had a higher risk of HMPV detection (AOR: 6.91, 95% CI: 1.71-47.3).

**Conclusion:** The study’s findings emphasize RSV infection in very young children and the potentially significant role of HMPV in causing respiratory infections among children in Ghana. Active surveillance of common respiratory viruses in healthcare facilities could enhance the management of viral respiratory infection cases in the paediatric population in Ghana.

## Introduction

Acute respiratory infections (ARIs) are important causes of illness worldwide, significantly burdening public health. They impact all age groups with a higher child mortality, especially in less developed countries [1]. Children in Sub-Saharan Africa are more than 15 times more likely to die before the age of five owing to ARIs [2]. ARIs in Ghana are part of the top 10 causes of hospital admissions and mortality in children under 5 years old [3].

Respiratory viruses account for a great percentage of ARIs. Respiratory viruses have been infecting humans for a long time and have evolved, leading to new strains and variants that cause different symptoms. The viruses that have been implicated as the etiological agents of ARIs include respiratory syncytial virus (RSV), influenza virus, human parainfluenza virus (HPIV), human rhinovirus (HRV), human adenovirus (HAdV) and human metapneumovirus (HMPV) [4]. RSV has been reported as the leading cause of severe lower respiratory tract infections with a burden of hospitalizations, especially in children in developing countries [5, 6, 7]. Pneumonia due to human metapneumovirus has shown a higher risk of intensive care need in Moroccan children [8]. Influenza virus is also reported as contributing substantially to the burden of hospitalized febrile children in Ghana [9]. The recent coronavirus disease 2019 (COVID-19) pandemic, caused by the severe acute respiratory syndrome coronavirus 2 (SARS-CoV-2), has further highlighted the role of respiratory viruses in ARIs.

The COVID-19 pandemic has profoundly reshaped the landscape of respiratory infections. During the pandemic, respiratory healthcare faced challenges, including disruptions to routine healthcare services, delays in immunization campaigns, and changes in healthcare-seeking patterns [10, 11]. These disruptions significantly influenced the prevalence and patterns of respiratory infections among children [12, 13,14]. As the acute phase of the pandemic caused significant shifts in societal norms, public health initiatives, and healthcare use trends, the transition to the post-pandemic era provides new challenges for healthcare systems and policymakers.

Despite the significant burden of respiratory viral infections among children in Ghana, there remains a paucity of comprehensive epidemiological data on circulating viruses with most studies skewed towards RSV and Influenza viruses. This study provides new insight into the changing dynamics of a broad spectrum of respiratory viruses among children in Ghana. The aim was to determine the prevalence of respiratory viruses and identify associated risk factors in children aged 5 or younger in an urban pediatric hospital setting.

## Materials and Methods

### Ethical consideration

The protocols used for the study were reviewed and approved by the Committee on Human Research and Publication Ethics (CHRPE), School of Medicine and Dentistry, Kwame Nkrumah University of Science and Technology (KNUST), Ghana (CHRPE/AP/135/22, CHRPE/AP/087/23) and parental consent was obtained from the caregivers/guardians of all the participants by signing or thumb-printing written consent forms.

### Study design and area

This was a cross-sectional hospital-based study conducted from August 2022 to June 2023 at the following facilities in Kumasi, Ashanti region of Ghana: The University Hospital, KNUST, Asokwa Children’s Hospital, Kumasi South Hospital, and HopeXchange Medical Centre. The KNUST hospital caters for about 150,000 people from over 30 surrounding communities. The Asokwa Children’s Hospital, which solely focuses on the care and management of children younger than 12 years, attends to roughly 6,000 children every month. The Kumasi South Hospital has an outpatient department and a paediatric ward that attends to approximately 4000 children monthly. The HopeXchange Medical Centre is a private facility with a paediatric ward that caters to about 1,500 children every month. All children 2 - 60 months who presented to the healthcare facilities with signs and symptoms of acute respiratory infections as defined in the WHO Programme for the Control of Acute Respiratory Infections [15] were enrolled. Participant selection was done without consideration of ethnicity and gender. Patients were excluded if they were over five years old, unable or unwilling to participate, or if they had any chronic lung disease. At enrolment, clinical symptoms and exposure were assessed, and a clinical examination was conducted to measure weight, height, respiratory rate, and peripheral capillary oxygen saturation.

### Data and sample collection

According to Obodai *et al* in 2019 [16], the prevalence of respiratory infections in Ghana was 23.0%. Considering this prevalence, the sample size was pre-determined to be 272 [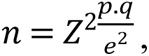 z = 1.9, p = 0.23, q = 0.77, e = 0.05]. A convenience sampling method was used throughout the study and patient recruitment was not done in parallel in each hospital. The general demographics, including age, sex, and maternal education, were recorded using a standardised questionnaire. Clinical data were extracted from the paediatrician’s folder. Clinical data included symptoms (cough, runny nose, headache, difficulty breathing, etc.) and diagnosis (URTI, bronchiolitis, pneumonia). Overall, 303 oropharyngeal swabs were collected with flocked swabs (Copan Group, Brescia, Italy) and inserted into 1.5mL Eppendorf tubes (Eppendorf, Germany) containing 500µl of viral transport medium (BioTeke Corporation Co., Ltd, China). The tubes were labelled with corresponding facility-specific identification numbers and transported on ice to the Kumasi Centre for Collaborative Research in Tropical Medicine (KCCR) for laboratory analysis.

### Laboratory Analysis

Nucleic acid extraction was done using the RADI PREP Swab and Stool DNA/RNA KIT (KH Medical Co., Ltd, Republic of Korea) according to the manufacturer’s instructions. Samples were eluted in a 70µl volume and SARS-CoV-2 viral RNA detection was performed using the RADI COVID-19 Detection Kit (KH Medical Co., Ltd, Republic of Korea) following the manufacturer’s instructions. The Invitrogen SuperScript III One-step RT-PCR System was used for the detection of Flu A, Flu B, RSV, PIV I-III, HRV and HMPV. For HAdV testing, the Qiagen 10X PCR buffer with Taq polymerase was used. Viral nucleic acid detection was done by real-time Polymerase Chain Reaction (PCR) using the LineGene 9600 Plus thermocycler (FQD-96A, Bioer Technology, China). All samples with a threshold cycle (Ct) above 40 were considered negative. Positive and negative controls were included in each PCR run for validation.

### Statistical Analysis

Data collected were entered into Microsoft Excel 2021, cleaned and exported to R programming language version 4.3.0 for analysis. Descriptive statistics were used to summarize the distribution of various variables into tables. Risk factors that had a significance level of p ≤ 0.05 in the univariate binary logistic regression analysis were subjected to multivariate binary logistic regression analysis. The results were expressed as the adjusted odds ratio (AOR) and 95% CI. All statistical tests with p-values less than 0.05 were considered statistically significant.

## Results

### Patient characteristics

From August 2022 to June 2023, a total of 303 children aged less than 5 years were recruited in this study. A majority (54.4%) were males aged 13 to 36 months (40.3%), with a median age of 19 months. The study participants presented with more than one symptom, with the most predominant being cough (87.0%), rhinorrhoea (87.0%) and fever (72.0%). The least recorded symptoms were headache (34.0%) and vomiting (25.0%). Table 1 shows the characteristics of the study population.

**Table 1:** Characteristics of the study population.

| Variable | Number of participants (%) |
| --- | --- |
| **Age (months)** |  |
| <6 | 45 (14.8) |
| 6-12 | 61 (20.1) |
| 13-36 | 122(40.3) |
| 37-60 | 75(24.8) |
| **Gender** |  |
| Male | 165 (54.4) |
| Female | 138 (45.6) |
| **Caregiver Education** |  |
| No formal education | 57 (19.0) |
| JHS/Primary | 42 (14.0) |
| SHS/Secondary | 54 (18.0) |
| Tertiary | 104 (34.0) |
| **Symptoms** |  |
| Fever | 219 (72.0) |
| Cough | 265 (87.0) |
| Headache | 102 (34.0) |
| Shortness of breath | 122 (40.0) |
| Poor feeding | 115 (38.0) |
| Vomiting | 76 (25.0) |
| Rhinorrhoea | 265 (87.0) |
| Loss of appetite | 117 (39.0) |

### Prevalence of respiratory viruses

Among the 303 samples tested, a respiratory virus was detected in 100 (33.0%) of the samples. In Table 2, it is shown that out of the samples tested, 92 (30.4%) showed single viral detections, while co-detections were observed in 8 samples (2.64%, 95% CI: 0.83-4.45). The most prevalent virus was HMPV with 36 cases (12.0%, 95% CI: 8.2-15.5), followed by RSV with 27 cases (8.9%, 95% CI: 5.7-12.1), and then HAdV with 20 cases (6.6%, 95% CI: 3.8-9.4). No PIV-I was detected.

**Table 2:** Prevalence of respiratory viruses.

| **Viral respiratory pathogen** | **Frequency** | **Prevalence (%)** |
| --- | --- | --- |
| SARS-CoV-2 | 3 | 1.0 |
| HAdV | 20 | 6.6 |
| HRV | 6 | 2.0 |
| PIV-I | 0 | 0 |
| PIV-II | 2 | 0.7 |
| PIV-III | 2 | 0.7 |
| FLU A | 5 | 1.7 |
| FLU B | 7 | 2.3 |
| HMPV | 36 | 12.0 |
| RSV | 27 | 8.9 |

The most predominant virus identified in cases of co-detection was HAdV (n=6, 75.0%). Table 3 shows that HMPV co-detection was identified in 4 (50.0%) samples. SARS-CoV-2, HRV, Flu A, Flu B, PIV-II, PIV-III and RSV were each identified in single cases of co-detection.

**Table 3:**
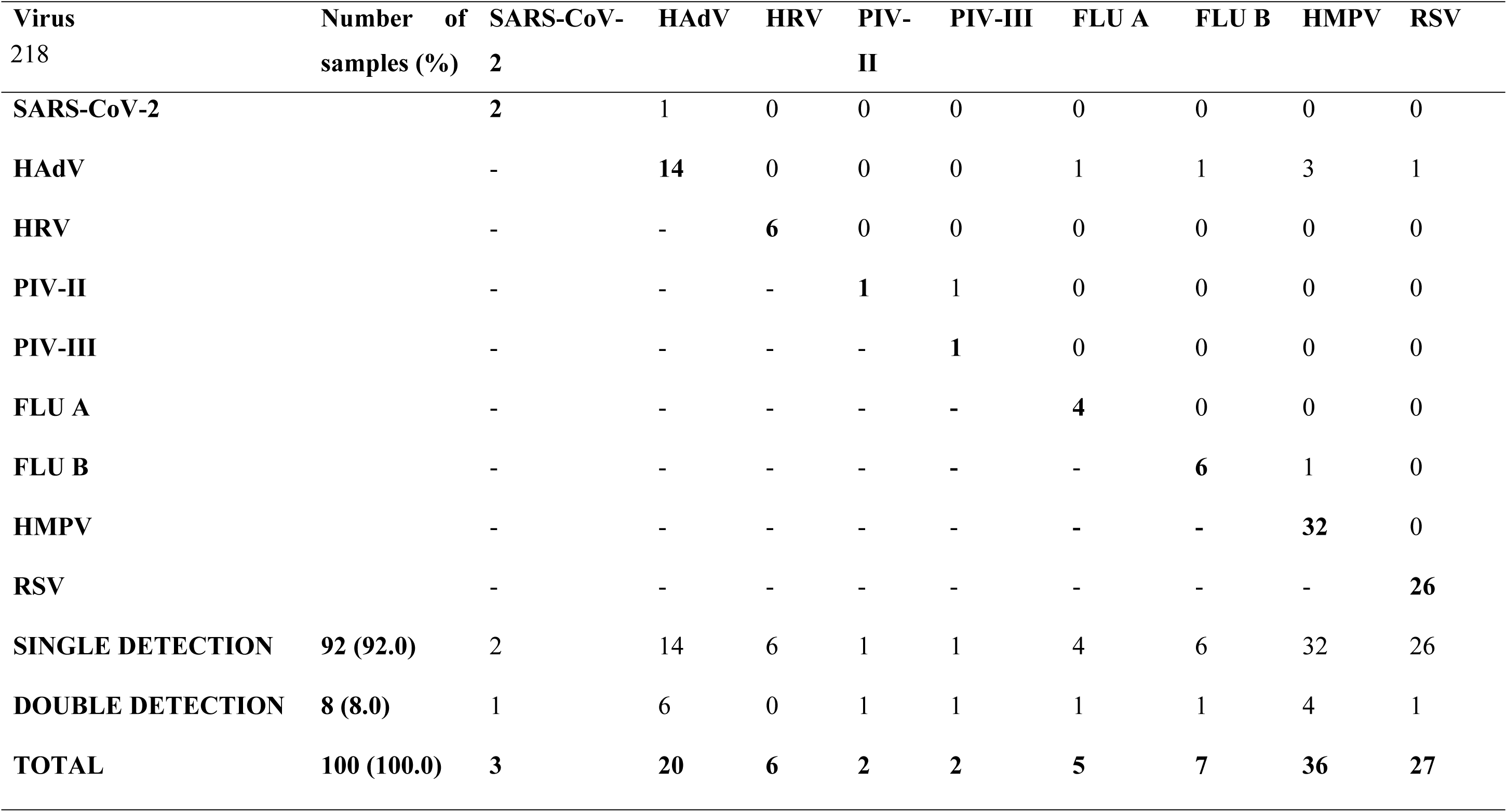
Single and double virus detections made during the study period.

### Risk factors associated with respiratory viral detection

The risk factor analysis was limited to HMPV, RSV, and HAdV due to the low prevalence of other respiratory viruses. According to the multivariate binary logistic regression analysis of the risk factors, children younger than 6 months (AOR: 4.81, 95% CI: 1.20-24.60) had higher odds of RSV detection when compared to children aged 37 to 60 months. It is evident from Table 4 that children with caregivers who had completed tertiary education (AOR: 6.91, 95% CI: 1.71-47.30) were more likely to test positive for HMPV compared to those with caregivers who had no formal education. The clinical risk factors evaluation in Table 5 showed that children presenting with headache (AOR: 3.95, 95% CI: 1.35-12.60) had higher odds of testing positive for HAdV compared to children without headache. In addition, children who presented with shortness of breath (AOR: 0.33, 95% CI: 0.13-1.02) had lower odds of testing positive for HMPV compared to children without shortness of breath.

**Table 4:** Risk factors associated with respiratory viruses in the study population.

| Variable | RSV | p-value | OR (95% CI) |  | AdV | p-value | RSV | p-value | AOR (95% CI) |  | AdV | p-value |
| --- | --- | --- | --- | --- | --- | --- | --- | --- | --- | --- | --- | --- |
|  |  |  | HMPV | p-value |  |  |  |  | HMPV | p-value |  |  |
| Age group |  |  |  |  |  |  |  |  |  |  |  |  |
| 37-60 | Ref |  | Ref |  | Ref |  | Ref |  | Ref |  | Ref |  |
| <6 | 6.00 | 0.010 | 0.60 | 0.500 | 0.53 | 0.500 | 4.81 | 0.036 | 0.79 | 0.700 | 0.55 | 0.500 |
|  | (1.68-28.30) |  | (0.13-2.20) |  | (0.08-2.44) |  | (1.20-24.60) |  | (0.16-3.11) |  | (0.07-2.71) |  |
| 6-12 | 2.14 | 0.300 | 0.91 | 0.900 | 0.59 | 0.500 | 2.21 | 0.300 | 0.96 | 0.900 | 0.60 | 0.500 |
|  | (0.50-10.80) |  | (0.29-2.78) |  | (0.12-2.36) |  | (0.48-11.90) |  | (0.29-3.06) |  | (0.12-2.48) |  |
| 13-36 | 2.14 | 0.300 | 1.54 | 0.300 | 0.92 | 0.900 | 2.27 | 0.200 | 1.39 | 0.500 | 0.91 | 0.900 |
|  | (0.63-9.8) |  | (0.66-3.93) |  | (0.32-2.84) |  | (0.64-10.70) |  | (0.57-3.62) |  | (0.31-2.86) |  |
| Gender |  |  |  |  |  |  |  |  |  |  |  |  |
| Female | Ref |  | Ref |  | Ref |  | Ref |  | Ref |  | Ref |  |
| Male | 0.46 | 0.062 | 0.82 | 0.600 | 0.83 | 0.700 | 0.46 | 0.069 | 0.89 | 0.800 | 0.83 | 0.700 |
|  | (0.20-1.02) |  | (0.40-1.65) |  | (0.33-2.07) |  | (0.19-1.05) |  | (0.41-1.96) |  | (0.31-2.18) |  |

| Caregiver |  |  |  |  |  |  |  |  |  |  |  |  |
| --- | --- | --- | --- | --- | --- | --- | --- | --- | --- | --- | --- | --- |
| Education |  |  |  |  |  |  |  |  |  |  |  |  |
| No education | Ref |  | Ref |  | Ref |  | Ref |  | Ref |  | Ref |  |
| JHS/Primary | 0.44 | 0.200 | 0.67 | 0.700 | 1.38 | 0.700 | - |  | 0.90 | 0.900 | 1.00 | 0.900 |
|  | (0.11-1.40) |  | (0.03-7.23) |  | (0.25-7.83) |  |  |  | (0.04-10.50) |  | (0.15-6.48) |  |
| SHS | - |  | 7.03 | **0.014** | 1.06 | 0.900 | - |  | 6.91 | **0.020** | 0.98 | 0.900 |
| /Secondary |  |  | (1.77-47.00) |  | (0.19-5.95) |  |  |  | (1.60-48.50) |  | (0.16-5.94) |  |
| Tertiary | 0.21 |  | 6.55 | **0.014** | 1.91 | 0.300 | - |  | 6.91 | **0.017** | 2.27 | 0.300 |
|  | (0.06-0.62) |  | (1.81-42.00) |  | (0.56-8.81) |  |  |  | (1.71-47.30) |  | (0.59) |  |

**Table 5:** Clinical symptoms associated with respiratory viruses in the study population.

| Variable | RSV | p-value | OR (95% CI) |  | AdV | p-value | RSV | p-value | AOR (95% CI) |  | AdV | p-value |
| --- | --- | --- | --- | --- | --- | --- | --- | --- | --- | --- | --- | --- |
|  |  |  | HMPV | p-value |  |  |  |  | HMPV | p-value |  |  |
| Clinical Symptom |  |  |  |  |  |  |  |  |  |  |  |  |
| Fever |  |  |  |  |  |  |  |  |  |  |  |  |
| No | Ref |  | Ref |  | Ref |  | Ref |  | Ref |  | Ref |  |
| Yes | 1.38 | 0.500 | 0.86 | 0.700 | 1.16 | 0.800 | 1.68 | 0.300 | 0.88 | 0.800 | 0.83 | 0.700 |
|  | (0.57-3.87) |  | (0.40-1.65) |  | (0.43-3.67) |  | (0.64-5.08) |  | (0.37-2.15) |  | (0.26-2.88) |  |
| Cough |  |  |  |  |  |  |  |  |  |  |  |  |
| No | Ref |  | Ref |  | Ref |  | Ref |  | Ref |  | Ref |  |
| Yes | - |  | 2.65 | 0.200 | 0.55 | 0.300 | - |  | 2.77 | 0.200 | 0.48 | 0.200 |
|  |  |  | (0.76-16.8) |  | (0.19-1.99) |  |  |  | (0.65-20.5) |  | (0.15-1.86) |  |
| Headache |  |  |  |  |  |  |  |  |  |  |  |  |
| No | Ref |  | Ref |  | Ref |  | Ref |  | Ref |  | Ref |  |
| Yes | 0.54 | 0.200 | 0.85 | 0.700 | 3.22 | **0.014** | 0.75 | 0.600 | 1.12 | 0.800 | 3.95 | **0.015** |
|  | (0.19-1.30) |  | (0.39-1.77) |  | (1.29-8.47) |  | (0.25-2.01) |  | (0.46-2.65) |  | (1.35-12.6) |  |
| **Shortness of breath** |  |  |  |  |  |  |  |  |  |  |  |  |
| No | Ref |  | Ref |  | Ref |  | Ref |  | Ref |  | Ref |  |
| Yes | 1.02 | 0.900 | 0.38 | **0.022** | 0.79 | 0.600 | 1.07 | 0.900 | 0.33 | **0.014** | 0.76 | 0.600 |
|  | (0.45-2.27) |  | (0.16-0.84) |  | (0.29-1.98) |  | (0.45-2.50) |  | (0.13-1.02) |  | (0.26-2.06) |  |
| **Rhinorrhoea** |  |  |  |  |  |  |  |  |  |  |  |  |
| No | Ref |  | Ref |  | Ref |  | Ref | 0.400 | Ref |  | Ref |  |
| Yes | 1.87 | 0.400 | 0.44 | 0.067 | - |  | 1.98 |  | 0.36 | 0.055 | - |  |
|  | (0.53-12.00) |  | (0.19-1.12) |  |  |  | (0.54-12.90) |  | (0.13-0.77) |  |  |  |
| **Poor feeding** |  |  |  |  |  |  |  |  |  |  |  |  |
| No | Ref |  | Ref |  | Ref |  | Ref |  | Ref |  | Ref |  |
| Yes | 0.80 | 0.600 | 0.43 | **0.043** | 1.37 | 0.500 | 0.81 | 0.600 | 0.39 | 0.052 | 1.07 | 0.900 |
|  | (0.33-1.81) |  | (0.18-0.93) |  | (0.53-3.41) |  | (0.32-1.94) |  | (0.15-0.97) |  | (0.38-2.94) |  |
| **Vomiting** |  |  |  |  |  |  |  |  |  |  |  |  |
| No | Ref |  | Ref |  | Ref |  | Ref |  | Ref |  | Ref |  |
| Yes | 0.84 | 0.700 | 0.34 | **0.048** | 2.11 | 0.120 | 0.81 | 0.700 | 0.55 | 0.300 | 2.13 | 0.140 |
|  | (0.30-2.05) |  | (0.10-0.89) |  | (0.80-5.31) |  | (0.27-2.11) |  | (0.15-1.64) |  | (0.75-5.84) |  |

## Discussion

The present study emphasized the prevalence of various respiratory viruses including RSV, Flu A, Flu B, HPIV I, II, III, HAdV, SARS-CoV-2, HRV, and HMPV among children exhibiting respiratory symptoms and signs. The overall prevalence recorded among the study participants was 33.0% (95% CI: 27.70-38.30). Previous studies from Ghana, prior to the COVID-19 pandemic, have reported varying estimates of prevalence of respiratory viruses from 12.5% to 73.0% among the paediatric population [1, 17, 18,19]. From those studies, relatively higher viral prevalence has been observed among hospitalized cases with more severe symptoms than cases recruited in the outpatient department (OPD), as was done in the present study [1, 17, 18,19]. In other parts of Africa, higher viral prevalence was reported in Burkina Faso (85.0%), Kenya (68.0%) and Cameroon (65.0%) [20–22]. Generally, variations in prevalence of respiratory viruses can be attributed to a number of reasons, including the panel of viral agents tested, the duration of the study, climatic conditions and enrolment criteria [23]. The recruitment of outpatients in the present study could account for the relatively lower viral prevalence in comparison with previously published studies that recruited hospitalized patients. HMPV was the most predominant virus, identified in 12.0% of the recruited participants. Approximately 90.0% of the HMPV cases were detected in one site during a three-week period, leading to a higher prevalence compared to previous reports in Ghana [1,18,19]. This increase may have been caused by an undetected HMPV outbreak, driven by nosocomial infections which is an existing significant public health concern, especially among paediatric patients [24]. The absence of viral testing in Ghanaian healthcare facilities further complicates the detection of such outbreaks. Recent studies since the decline of SARS-CoV-2 cases have also reported high prevalence of HMPV in Myanmar (13.0%), USA (12.6%) and Nepal (13.3%) [5,25,26]. RSV, which has been implicated as a major cause of ARI in children, was identified in 8.9% of the study population. The aggressive implementation of non-pharmaceutical interventions (NPIs) during the COVID-19 pandemic led to a significant decline in RSV cases among young children globally, as reported in South Africa (4.1%), the United Kingdom (0.1%) and Canada (0.05%) [12,13,27]. However, with the gradual relaxation of NPIs, countries like Egypt (20.9%) and China (21.7%) witnessed a resurgence in RSV cases, particularly among young children [28,29]. The finding from this study emphasizes the importance of active surveillance in enhancing the management of RSV-related respiratory infections in young children. The prevalence of HAdV (6.6%), HRV (2.0%), flu A (1.7%), and Flu B (2.3%) in the present study was lower than previous studies conducted pre-COVID-19 pandemic among Ghanaian children [9,17–19]. Apart from Influenza, which is under routine surveillance in Ghana, respiratory viruses are not routinely sought for in Ghanaian healthcare facilities [(9)]. The relatively lower prevalence still, however, highlights the circulation of these viruses and a major implication of the failure to detect them may cause an increase in antimicrobial resistance as antibiotics are often prescribed to treat apparent respiratory infections [30]. Young children have been reported to be less often infected by SARS-CoV-2 when compared to adults [31], and unsurprisingly, the 1.0% prevalence from our study was similarly as low as reported among children (0.4%) during the peak of the COVID-19 pandemic in Ghana [32]. Comparisons with studies from other parts of the world revealed that regardless of the clinical profile, SARS-CoV-2 prevalence remains low among children [33–35]. HPIV-II and HPIV-III were both detected in 0.7% of the study group with no detection (0%) for HPIV-I. This was lower than previous reports in Ghana [36, 17,19], and other African countries like Cameroon and Niger [22,23]. It is possible there may have been low viral activity of HPIV since there was no year-round sampling. Generally, the limited diagnostic capacity of healthcare facilities to detect respiratory viruses increases the risk of infected children unknowingly transmitting these viruses to others in daycare, schools and households. Consequently, this adds more burden to the constrained healthcare system in Ghana.

HMPV and HAdV accounted for 3 (37.5%) of the 8 co-detections observed in this study. Single cases of co-detection were observed for the HAdV and RSV, HAdV and SARS-CoV-2, HAdV and flu A, HMPV and flu B, and PIV-II and PIV-III. Literature on viral co-detection in respiratory illness among children shows conflicting results. Some studies [37,38] suggest worse outcomes and longer hospital stays, whereas others [39,40] found no association between viral co-detection and illness severity. Our study however, did not determine the association between viral co-detection and its impact on health, as only outpatients were recruited.

We identified age as a risk factor for RSV detection in our study. Children aged younger than 6 months (AOR: 4.81, 95% CI: 1.20-24.60) had higher odds of testing positive for RSV when compared to those aged from 37 to 60 months. This is consistent with other studies that identified a higher risk of RSV infection among children less than 6 months than other age groups [41,42]. Children younger than six months are biologically more vulnerable to ARIs due to the ongoing development of their immune system and lungs, leaving them with insufficient immunity when exposed to infections [43].

Our study identified secondary and tertiary level of caregiver education as risk factors for HMPV detection. Children whose caregivers had attained secondary (AOR: 6.91, 95% CI: 1.60-48.50) and tertiary (AOR: 6.91, 95% CI: 1.71-47.30) education had higher odds of HMPV detection when compared to those with caregivers of no formal education. This was in contrast to findings by Ujunwa in Nigeria and Tazinya in Cameroon, who suggested that no formal education among caregivers led to poor hygienic practices in homes which increased the risk of respiratory viral detection among children [44,45]. A possible explanation for our finding could be that caregivers with higher education may enrol children in crowded daycare settings and kindergartens, potentially increasing the risk of respiratory virus transmission, such as HMPV. We identified headache as a clinical risk factor for HAdV detection in our study.

Children with headaches (AOR: 3.95, 95% CI: 1.35-12.60) had higher odds of testing positive for HAdV than those without headaches. This aligns with a study in Senegal linking HAdV detection to headaches in children with respiratory illness [46]. Headaches in infants have been associated with adenoviral pharyngitis, and common cold symptoms induced by HAdV, such as sneezing and sinus inflammation, which can contribute to headache sensations [47,48]. Children with shortness of breath (AOR: 0.33, 95% CI: 0.13-1.02) in our study had lower odds of testing positive for HMPV, contrary to previous studies associating dyspnoea with HMPV infection [49–51]. The discrepancy might be due to potential non-viral causes, like *Streptococcus pneumoniae* or *Haemophilus influenzae*, which were not assessed in this study but are known to cause bacterial pneumonia with similar symptoms [52].

## Conclusion

The results of our study indicate that HMPV, the most common respiratory virus, may have a more significant impact on the cause of respiratory illness in Ghanaian children than previously believed. The increased scientific and public awareness of the burden of respiratory infections by the COVID-19 pandemic provides a good opportunity to make advances towards reducing the spread of common respiratory viruses. Further studies should be conducted in other geographical locations in the country to elucidate the viral prevalence patterns and provide a comprehensive overview of the respiratory virome in the Ghanaian paediatric population.

## Data Availability

All relevant data are within the manuscript (Accession number: DOI 10.17605/OSF.IO/QW2HU

## Acknowledgements

We want to acknowledge all participants and their families who agreed to participate in the study. Additionally, we also acknowledge Annie Sarpong Animwaa and Abigail Achiaa Akoto for their help in participant recruitment. There was no specific funding for this study.

## Supporting information

S1_Checklist: Clinical Studies Checklist

S2_Checklist: STROBE checklist

**S3 data**. Dataset for study: DOI 10.17605/OSF.IO/QW2HU

## References

1. Kafintu-Kwashie AA, Nii-Trebi NI, Obodai E, Neizer M, Adiku TK, Odoom JK. Molecular epidemiological surveillance of viral agents of acute lower respiratory tract infections in children in Accra, Ghana. BMC Pediatr [Internet]. 2022;22(1):1–9. Available from: 10.1186/s12887-022-03419-7

2. Apanga PA, Kumbeni MT. Factors associated with diarrhoea and acute respiratory infection in children under-5 years old in Ghana: an analysis of a national cross-sectional survey. BMC Pediatr. 2021;21(1):1–8.

3. Abbey M, Afagbedzi SK, Afriyie-Mensah J, Antwi-Agyei D, Atengble K, Badoe E, et al. Pneumonia in Ghana-a need to raise the profile. Int Health. 2018;10(1):4–7.

4. Falkenstein-Hagander K, Mansson AS, Redmo J, Nilsson Wimar P, Widell A. Viral aetiology and clinical outcomes in hospitalised infants presenting with respiratory distress. Acta Paediatr Int J Paediatr. 2014;103(6):625–9.

5. Lamichhane J, Upreti M, Nepal K, Upadhyay BP, Maharjan U, Shrestha RK, et al. Burden of human metapneumovirus infections among children with acute respiratory tract infections attending a Tertiary Care Hospital, Kathmandu. BMC Pediatr [Internet]. 2023 Dec 1 [cited 2023 Nov 12];23(1). Available from: /pmc/articles/PMC10405573/

6. Shafik CF, Mohareb EW, Yassin AS, Amin MA, El Kholy A, El-Karaksy H, et al. Viral etiologies of lower respiratory tract infections among Egyptian children under five years of age. BMC Infect Dis [Internet]. 2012 Dec 13 [cited 2023 Sep 15];12(1):1–8. Available from: https://bmcinfectdis.biomedcentral.com/articles/10.1186/1471-2334-12-350

7. O’Callaghan-Gordo C, Díez-Padrisa N, Abacassamo F, Pérez-Breña P, Casas I, Alonso PL, et al. Viral acute respiratory infections among infants visited in a rural hospital of southern Mozambique. Trop Med Int Heal [Internet]. 2011 Sep 1 [cited 2023 Sep 25];16(9):1054–60. Available from: https://onlinelibrary.wiley.com/doi/full/10.1111/j.1365-3156.2011.02811.x

8. Jroundi I, Mahraoui C, Benmessaoud R, Moraleda C, Tligui H, Seffar M, et al. A comparison of human metapneumovirus and respiratory syncytial virus WHO-defined severe pneumonia in Moroccan children. Epidemiol Infect [Internet]. 2016 Feb 1 [cited 2024 May 6];144(3):516–26. Available from: https://www.cambridge.org/core/journals/epidemiology-and-infection/article/comparison-of-human-metapneumovirus-and-respiratory-syncytial-virus-whodefined-severe-pneumonia-in-moroccan-children/84C1BCBF6A98FFDA3BB9CFAE27C63222

9. Hogan B, Ammer L, Zimmermann M, Binger T, Krumkamp R, Sarpong N, et al. Burden of influenza among hospitalized febrile children in Ghana. Influenza Other Respi Viruses. 2017;11(6):497–501.

10. Afulani PA, Gyamerah AO, Nutor JJ, Laar A, Aborigo RA, Malechi H, et al. Inadequate preparedness for response to COVID-19 is associated with stress and burnout among healthcare workers in Ghana. PLoS One [Internet]. 2021 Apr 1 [cited 2023 Mar 29];16(4):e0250294. Available from: https://journals.plos.org/plosone/article?id=10.1371/journal.pone.0250294

11. Rehman SU, Rehman SU, Yoo HH. COVID-19 challenges and its therapeutics. Biomed Pharmacother [Internet]. 2021 Oct 1 [cited 2024 May 6];142:112015. Available from: /pmc/articles/PMC8339548/

12. Tempia S, Walaza S, Bhiman JN, McMorrow ML, Moyes J, Mkhencele T, et al. Decline of influenza and respiratory syncytial virus detection in facility-based surveillance during the COVID-19 pandemic, South Africa, January to October 2020. Eurosurveillance [Internet]. 2021 Jul 7 [cited 2023 Nov 13];26(29). Available from: /pmc/articles/PMC8299743/

13. Groves HE, Piché-Renaud PP, Peci A, Farrar DS, Buckrell S, Bancej C, et al. The impact of the COVID-19 pandemic on influenza, respiratory syncytial virus, and other seasonal respiratory virus circulation in Canada: A population-based study. Lancet Reg Heal - Am [Internet]. 2021 Sep 1 [cited 2023 Mar 19];1:15. Available from: /pmc/articles/PMC8285668/

14. Kandeel A, Fahim M, Deghedy O, Roshdy WH, Khalifa MK, Shesheny R El, et al. Resurgence of influenza and respiratory syncytial virus in Egypt following two years of decline during the COVID-19 pandemic: outpatient clinic survey of infants and children, October 2022. BMC Public Health [Internet]. 2023 Dec 1 [cited 2023 Nov 13];23(1):1–9. Available from: https://bmcpublichealth.biomedcentral.com/articles/10.1186/s12889-023-15880-9

15. World Health Organization. Acute respiratory infections in children: Case management in small hospitals in developing countries. 1990.

16. Obodai E, Odoom JK, Adiku T, Goka B, Wolff T, Biere B, et al. Erratum: The significance of human respiratory syncytial virus (HRSV) in children from Ghana with acute lower respiratory tract infection: A molecular epidemiological analysis, 2006 and 2013-2014 (PLoS ONE (2019) 13:9 (e0203788)Doi: 10.1371/journal.pone.0. PLoS One. 2019;14(8):1–17.

17. Annan A, Ebach F, Corman VM, Krumkamp R, Adu-Sarkodie Y, Eis-Hübinger AM, et al. Similar virus spectra and seasonality in paediatric patients with acute respiratory disease, Ghana and Germany. Clin Microbiol Infect [Internet]. 2016 Apr 1 [cited 2023 May 17];22(4):340. Available from: /pmc/articles/PMC7172147/

18. Krumkamp R, Kohsar M, Nolte K, Hogan B, Eibach D, Jaeger A, et al. Pathogens associated with hospitalization due to acute lower respiratory tract infections in children in rural Ghana: a case-control study. Sci Rep [Internet]. 2023;13(1):2443. Available from: 10.1038/s41598-023-29410-5

19. Obodai E. Molecular Epidemiology of Respiratory Viruses associated with Acute Lower Respiratory Tract Infections in Children from Ghana. 2016; Available from: https://refubium.fu-berlin.de/handle/fub188/2268

20. Ouédraogo S, Traoré B, Bi ZABN, Yonli FT, Kima D, Bonané P, et al. Viral etiology of respiratory tract infections in children at the pediatric hospital in Ouagadougou (Burkina Faso). PLoS One. 2014 Oct 31;9(10).

21. Feikin DR, Njenga MK, Bigogo G, Aura B, Aol G, Audi A, et al. Viral and bacterial causes of severe acute respiratory illness among children aged less than 5 years in a high malaria prevalence area of Western Kenya, 2007-2010. Pediatr Infect Dis J [Internet]. 2013 Jan [cited 2023 Sep 7];32(1). Available from: https://journals.lww.com/pidj/fulltext/2013/01000/viral_and_bacterial_causes_of_severe_acute.10.aspx

22. Kenmoe S, Tchendjou P, Vernet MA, Moyo-Tetang S, Mossus T, Njankouo-Ripa M, et al. Viral etiology of severe acute respiratory infections in hospitalized children in Cameroon, 2011–2013. Influenza Other Respi Viruses [Internet]. 2016 Sep 1 [cited 2023 Sep 8];10(5):386. Available from: /pmc/articles/PMC4947949/

23. Lagare A, Maïnassara HB, Issaka B, Sidiki A, Tempia S. Viral and bacterial etiology of severe acute respiratory illness among children < 5 years of age without influenza in Niger. BMC Infect Dis [Internet]. 2015 Nov 14 [cited 2023 Sep 8];15(1):1–7. Available from: https://bmcinfectdis.biomedcentral.com/articles/10.1186/s12879-015-1251-y

24. Chow EJ, Mermel LA. Hospital-Acquired Respiratory Viral Infections: Incidence, Morbidity, and Mortality in Pediatric and Adult Patients. Open Forum Infect Dis [Internet]. 2017 Jan 1 [cited 2023 Sep 15];4(1). Available from: /pmc/articles/PMC5414085/

25. Kamata K, Thein KN, Di Ja L, Win NC, Win SMK, Suzuki Y, et al. Clinical manifestations and outcome of viral acute lower respiratory infection in hospitalised children in Myanmar. BMC Infect Dis [Internet]. 2022 Dec 1 [cited 2023 Sep 28];22(1):1–17. Available from: https://bmcinfectdis.biomedcentral.com/articles/10.1186/s12879-022-07342-1

26. Howard LM, Edwards KM, Zhu Y, Grijalva CG, Self WH, Jain S, et al. Clinical Features of Human Metapneumovirus-Associated Community-acquired Pneumonia Hospitalizations. Clin Infect Dis An Off Publ Infect Dis Soc Am [Internet]. 2021 Jan 1 [cited 2023 Oct 5];72(1):108. Available from: /pmc/articles/PMC7823075/

27. Bardsley M, Morbey RA, Hughes HE, Beck CR, Watson CH, Zhao H, et al. Epidemiology of respiratory syncytial virus in children younger than 5 years in England during the COVID-19 pandemic, measured by laboratory, clinical, and syndromic surveillance: a retrospective observational study. Lancet Infect Dis [Internet]. 2023 Jan 1 [cited 2023 Nov 13];23(1):56. Available from: /pmc/articles/PMC9762748/

28. Abu-Raya B, Viñeta Paramo M, Reicherz F, Lavoie PM. Why has the epidemiology of RSV changed during the COVID-19 pandemic? eClinicalMedicine [Internet]. 2023 Jul 1 [cited 2023 Nov 13];61. Available from: http://www.thelancet.com/article/S2589537023002663/fulltext

29. Chuang YC, Lin KP, Wang LA, Yeh TK, Liu PY. The Impact of the COVID-19 Pandemic on Respiratory Syncytial Virus Infection: A Narrative Review. Infect Drug Resist [Internet]. 2023 [cited 2023 Nov 13];16:661. Available from: /pmc/articles/PMC9897071/

30. Labi AK, Obeng-Nkrumah N, Nartey ET, Bjerrum S, Adu-Aryee NA, Ofori-Adjei YA, et al. Antibiotic use in a tertiary healthcare facility in Ghana: A point prevalence survey. Antimicrob Resist Infect Control [Internet]. 2018 Jan 26 [cited 2023 Nov 12];7(1):1–9. Available from: https://aricjournal.biomedcentral.com/articles/10.1186/s13756-018-0299-z

31. Howard-Jones AR, Bowen AC, Danchin M, Koirala A, Sharma K, Yeoh DK, et al. COVID-19 in children: I. Epidemiology, prevention and indirect impacts. J Paediatr Child Health [Internet]. 2022 Jan 1 [cited 2023 Sep 26];58(1):39. Available from: /pmc/articles/PMC8662210/

32. Owusu M, Sylverken AA, Ankrah ST, El-Duah P, Ayisi-Boateng NK, Yeboah R, et al. Epidemiological profile of SARS-CoV-2 among selected regions in Ghana: A cross-sectional retrospective study. PLoS One. 2020;15(12 December):1–15.

33. Hua W, Xiaofeng L, Zhenqiang B, Jun R, Ban W, Liming L. [The epidemiological characteristics of an outbreak of 2019 novel coronavirus diseases (COVID-19) in China]. Zhonghua Liu Xing Bing Xue Za Zhi [Internet]. 2020 Feb 1 [cited 2023 Sep 26];41(2):297–300. Available from: https://pubmed.ncbi.nlm.nih.gov/32064853/

34. Wu Z, McGoogan JM. Characteristics of and Important Lessons From the Coronavirus Disease 2019 (COVID-19) Outbreak in China: Summary of a Report of 72 314 Cases From the Chinese Center for Disease Control and Prevention. JAMA [Internet]. 2020 Apr 7 [cited 2023 Jun 11];323(13):1239–42. Available from: https://jamanetwork.com/journals/jama/fullarticle/2762130

35. Eskander E, Jung C, Levy C, Béchet S, Blot N, Gorde S, et al. Assessment of SARS-CoV-2 testing in children during a low prevalence period (VIGIL study 1). Infect Dis Now [Internet]. 2021 Sep 1 [cited 2023 Sep 26];51(6):552. Available from: /pmc/articles/PMC8276575/

36. Kwofie TB, Anane YA, Nkrumah B, Annan A, Nguah SB, Owusu M. Respiratory viruses in children hospitalized for acute lower respiratory tract infection in Ghana. Virol J [Internet]. 2012 Apr 10 [cited 2023 May 17];9(1):1–8. Available from: https://virologyj.biomedcentral.com/articles/10.1186/1743-422X-9-78

37. Xie L, Zhang B, Xiao N, Zhang F, Zhao X, Liu Q, et al. Epidemiology of human adenovirus infection in children hospitalized with lower respiratory tract infections in Hunan, China. J Med Virol [Internet]. 2019 Mar 1 [cited 2023 Jun 10];91(3):392. Available from: /pmc/articles/PMC7159165/

38. Arnold FW, Fuqua JL. Viral respiratory infections: A cause of community-acquired pneumonia or a predisposing factor? Curr Opin Pulm Med [Internet]. 2020 May 1 [cited 2023 May 24];26(3):208–14. Available from: https://journals.lww.com/co-pulmonarymedicine/Fulltext/2020/05000/Viral_respiratory_infections__a_cause_of.5.aspx

39. Scotta MC, Chakr VCBG, de Moura A, Becker RG, de Souza APD, Jones MH, et al. Respiratory viral coinfection and disease severity in children: A systematic review and meta-analysis. J Clin Virol [Internet]. 2016 Jul 1 [cited 2023 May 24];80:45. Available from: /pmc/articles/PMC7185664/

40. Swets MC, Russell CD, Harrison EM, Docherty AB, Lone N, Girvan M, et al. SARS-CoV-2 co-infection with influenza viruses, respiratory syncytial virus, or adenoviruses. Lancet (London, England) [Internet]. 2022 Apr 16 [cited 2023 May 24];399(10334):1463. Available from: /pmc/articles/PMC8956294/

41. Hasan MM, Saha KK, Yunus RM, Alam K. Prevalence of acute respiratory infections among children in India: Regional inequalities and risk factors. Matern Child Health J [Internet]. 2022 Jul 1 [cited 2024 May 6];26(7):1594. Available from: /pmc/articles/PMC9174316/

42. Sun YP, Qiang HS, Lei SY, Zheng XY, Zhang HX, Su YY, et al. Epidemiological Features, Risk Factors, and Disease Burden of Respiratory Viruses among Hospitalized Children with Acute Respiratory Tract Infections in Xiamen, China. Jpn J Infect Dis [Internet]. 2022 [cited 2024 May 6];75(6):537–42. Available from: https://pubmed.ncbi.nlm.nih.gov/35768274/

43. Schraufnagel DE, Balmes JR, Cowl CT, De Matteis S, Jung SH, Mortimer K, et al. Air Pollution and Noncommunicable Diseases: A Review by the Forum of International Respiratory Societies’ Environmental Committee, Part 2: Air Pollution and Organ Systems. Chest [Internet]. 2019 Feb 1 [cited 2024 May 6];155(2):417. Available from: /pmc/articles/PMC6904854/

44. Ujunwa F, Ezeonu C. Risk Factors for Acute Respiratory Tract Infections in Under-five Children in Enugu Southeast Nigeria. Ann Med Health Sci Res [Internet]. 2014 [cited 2023 May 27];4(1):95. Available from: /pmc/articles/PMC3952306/

45. Tazinya AA, Halle-Ekane GE, Mbuagbaw LT, Abanda M, Atashili J, Obama MT. Risk factors for acute respiratory infections in children under five years attending the Bamenda Regional Hospital in Cameroon. BMC Pulm Med [Internet]. 2018 Jan 16 [cited 2023 Oct 4];18(1). Available from: /pmc/articles/PMC5771025/

46. Niang MN, Diop NS, Fall A, Kiori DE, Sarr FD, Sy S, et al. Respiratory viruses in patients with influenza-like illness in Senegal: Focus on human respiratory adenoviruses. PLoS One [Internet]. 2017 Mar 1 [cited 2023 Oct 5];12(3). Available from: /pmc/articles/PMC5362214/

47. Shieh WJ. Human adenovirus infections in pediatric population - An update on clinico– pathologic correlation. Biomed J [Internet]. 2022 Feb 1 [cited 2023 Oct 5];45(1):38. Available from: /pmc/articles/PMC9133246/

48. Kirkpatrick GL. THE COMMON COLD. Prim Care [Internet]. 1996 Dec 12 [cited 2023 Oct 5];23(4):657. Available from: /pmc/articles/PMC7125839/

49. Williams J V., Edwards KM, Weinberg GA, Griffin MR, Hall CB, Zhu Y, et al. Population-Based Incidence of Human Metapneumovirus Infection among Hospitalized Children. J Infect Dis [Internet]. 2010 Jun 15 [cited 2023 Oct 5];201(12):1890–8. Available from: 10.1086/652782

50. Howard LM, Edwards KM, Zhu Y, Griffin MR, Weinberg GA, Szilagyi PG, et al. Clinical Features of Human Metapneumovirus Infection in Ambulatory Children Aged 5–13 Years. J Pediatric Infect Dis Soc [Internet]. 2018 [cited 2023 Oct 5];7(2):165. Available from: /pmc/articles/PMC5954304/

51. Noordeen F, Pitchai FNN, Kudagammana ST, Rafeek RAM. A mini outbreak of human metapneumovirus infection with severe acute respiratory symptoms in a selected group of children presented to a teaching hospital in Sri Lanka. VirusDisease [Internet]. 2019 Jun 1 [cited 2023 Oct 5];30(2):307–10. Available from: https://link.springer.com/article/10.1007/s13337-019-00522-9

52. Nathan AM, Teh CSJ, Jabar KA, Teoh BT, Tangaperumal A, Westerhout C, et al. Bacterial pneumonia and its associated factors in children from a developing country: A prospective cohort study. PLoS One [Internet]. 2020 Feb 1 [cited 2023 Oct 5];15(2). Available from: /pmc/articles/PMC7021284/

